# Changes in somatic symptoms among people with severe COVID anxiety, before and after the coronavirus pandemic

**DOI:** 10.1101/2025.02.26.25322939

**Authors:** Leila Grocott, Jacob D. King, Aisling McQuaid, Verity C. Leeson, Mike J. Crawford

## Abstract

**Background:** Anxiety and somatic symptoms were common during the coronavirus pandemic. People who were highly anxious about COVID-19 may have been at a higher risk of developing new somatic symptoms which persisted after the pandemic. Here we examine changes in somatic symptoms before and after the pandemic among people who had severe COVID anxiety, and identify factors associated with these changes.

**Design:** UK adults who met the threshold for severe COVID anxiety were recruited online during the coronavirus pandemic, and were asked to rate their somatic symptoms post-pandemic, and retrospectively before the pandemic with the PHQ-15. Data on demographic and clinical factors were also collected.

**Methods:** Descriptive statistics and multiple linear regression were used to characterise the sample of 197 people who provided complete data, and to examine factors associated with changes in somatic symptoms.

**Results:** Mean PHQ-15 score increased significantly from 7.97 before the pandemic to 11.34 afterwards (t= 9.043, p<0.001). Despite this, there were gradual corresponding declines in levels of COVID anxiety and other co-occurring mental health symptoms after the pandemic ended. Multiple linear regression models identified that greater generalised anxiety symptoms and living with someone vulnerable to COVID were the strongest predictors of increased somatic symptoms during the pandemic.

**Conclusions:** People with severe COVID anxiety reported high rates of somatic symptoms before the pandemic, which then increased significantly after the pandemic. There appears to be a complex interaction between COVID anxiety, pandemic experience, and somatic symptoms which warrants further investigation, and could inform targeted intervention in the future.

**What is already known on this subject?:**

- A host of demographic and psychological factors, such as generalised and health anxiety, contribute to the reporting of somatic symptoms.
- Recent evidence suggests that individuals with COVID-19 specific anxiety are more likely to report somatic symptoms, and that this association was present after controlling for other demographic and psychological factors.
- People who lived with someone vulnerable to COVID-19 during the pandemic reported higher rates of pandemic related stress and mental health symptoms.

**What does this study add?:**

- Individuals with severe levels of COVID anxiety show a higher somatic symptom count compared to the general population.
- Chest pain, dizziness and palpitations were the somatic symptoms which were most frequently reported to have increased over the pandemic by people who had severe levels of COVID anxiety.
- The combination of severe COVID anxiety and living with someone vulnerable to COVID appears to have contributed to a higher risk of worsening somatic symptoms during the pandemic.

## Introduction

Somatic symptoms are bodily sensations that cause discomfort and distress^1^ and the more somatic symptoms a person has, the worse their mental and physical health tends to be^2^. There is a close association between reporting somatic symptoms and anxiety^3^, which are thought by some to be connected by both of their links to physical illness^4, 5^ or indeed shared processes of hypervigilance^6^, and biases in attention and predictive coding^7^, among others.

Anxious thoughts and feelings about COVID-19 were appropriately common during the pandemic, but for a small minority this level of preoccupation, and concerns with contagion and the health and socioeconomic consequences of the pandemic, became severe^8^. For some people physical symptoms of anxiety like shortness of breath, gastrointestinal disturbance and palpitations were believed to be evidence of COVID-19 infection, reinforcing a preoccupation with COVID-19^9^. This could lead to the use of health behaviours intended to have a protective effect, like washing behaviours, strict isolation, and symptom-checking behaviours, but which could be performed to a degree in excess of any public health guidelines, amplifying distress and preventing people from doing the other important things in their lives.

Concerns were raised at the start of the COVID-19 pandemic about the impact the spread of the virus might have on population mental health, and on the prevalence of somatic symptoms in particular^10, 11^. A systematic review exploring somatic symptoms throughout the COVID-19 pandemic suggested that up to 63% of the general population in Brazil and 54% in Poland experienced somatic symptoms in the period from April to July 2020^12^. Indeed, evidence from a population-based study in the UK with over 2,000 participants demonstrated that the number and impact of somatic symptoms was positively associated with the level of COVID anxiety^10^.

Moreover, several studies have examined changes in somatic symptoms over the course of the pandemic, demonstrating sizeable increases among Norwegian adolescents^13^ and Turkish adults^14^. From the limited number of demographic and clinical factors these studies collected, female gender, being a healthcare worker, perception of threat, and having contracted COVID appeared associated with greater increases in somatic symptoms over the pandemic^12-14^.

The present study aims to further prospect a link between COVID anxiety and somatic symptoms^15^ using a wide selection of demographic and clinical factors. A more complete understanding of this relationship, particularly focused on those with severe levels of COVID anxiety, might better place clinicians to assess and support these experiences where appropriate. It was hypothesised that among people with severe COVID anxiety, somatic symptoms would increase throughout the pandemic and that higher levels of somatic symptoms would be associated with a persistence of high levels of generalised anxiety, health anxiety and COVID anxiety over time.

## Methods

This is a secondary analysis of data from the COVID Anxiety Project^16^ which investigated the mental health of people with severe COVID anxiety longitudinally over an 18 month follow-up period. The study was pre-registered (ISRCTN14973494), and a protocol is previously published^15^.

### Recruitment

People who self-identified as being anxious about COVID were recruited via adverts on social media and from 19 general practice clinics in North West London between February 2021 and September 2021^16^. This was at a point of the pandemic in the UK more than a year after the first cases had been reported, and over which the third national lockdown was in effect, then ended, and distancing measures were gradually easing. Participants were eligible for the cohort if they 1) scored 9 or more on the Coronavirus Anxiety Scale (CAS) indicating a severe impact of COVID anxiety on daily life, 2) were aged ≥ 18 years, 3) a resident of the UK, 4) have sufficient ability in English to participate, and 5) reported no history of psychosis^16, 17^. The participants were followed up with further online surveys at 3 months, 6 months, and 18 months, with the last datapoint collected in April 2023. During the 18-month collection period there were no public heath restrictions in place in UK and no national case detection efforts. The surveys were created and distributed using the Qualtrics online application^15^.

### Ethics

Leicester Central Research Ethics Committee and Health Regulation Authority approved the COVID Anxiety Project, reference number 20/EM/023^16^. The study adhered to the guidelines established by the Declaration of Helsinki 1964^16^. Data were handled according to the General Data Protection regulations, and Qualtrics is certified under ISO 27001, ensuring it adheres to international standards for handling sensitive and confidential information^15^. All participants signed a consent form before taking part in the study. Once completed, the consent forms were printed and physically filed securely^15^. Participants were offered a £10 gift voucher for completing the baseline survey and £20 gift vouchers for completing the 6-month survey, and 18-month surveys. Participants could withdraw from the study at any point with no explanation required.

### Measurements

#### Primary outcome

At the 18-month follow-up point, participants completed a battery of questionnaires that included the PHQ-15, a 15-item scale assessing somatic symptoms. Respondents rate 15 symptoms, such as back pain and shortness of breath, in the preceding 4 weeks, on a scale from 0 (not bothered at all by the symptom) to 2 (bothered a lot)^2^. They were asked to complete both a PHQ-15 for any somatic symptoms they were currently experiencing, and then retrospectively a PHQ-15, covering somatic symptoms they recalled experiencing before the start of the pandemic.

The PHQ-15 is a widely used and validated self-report questionnaire that measures the severity of somatic symptoms^2^, with greater scores representing a greater number of, and more severe, somatic symptoms. Increasing PHQ-15 scores are negatively correlated with functional status^2^. The maximum score is 30, and severity thresholds in the literature are as follows: low (5-9), medium (10-14), high (15-30)^2^. Sensitivity to detecting somatoform disorders at a cut-off of ≥ 3 severe somatic symptoms is reported to be 0.78, with a specificity of 0.71^18^.

### Mental health measures

The following self-report questionnaires were also completed at baseline and repeated at 18 months.

The Coronavirus Anxiety Scale (CAS) is a validated self-report questionnaire that assesses the severity of 5 psychophysiological symptoms that might arise when exposed to information about the COVID-19 pandemic^17, 19^. It was the only scale validated for assessing COVID anxiety at the beginning of the COVID Anxiety Project^15^. The scale has a maximum score of 20 (maximum rating of 4 for each question), and a score of 9 or above is associated with significant impairment in daily function and severe COVID anxiety^17, 19^.

In addition, the 7-item Generalised Anxiety Disorder Scale (GAD-7), the 14-item Short Health Anxiety Inventory (sHAI -14), the 9-item Patient Health Questionnaire (PHQ-9), and the 3-item Alcohol Use Disorders Identification Test – Consumption (AUDIT-C) were collected. These are self-report scales widely used for briefly assessing symptoms of generalised anxiety, health anxiety, depression, and hazardous alcohol use respectively, validated within both clinical and general populations^20-23^.

The Obsessive-Compulsive Inventory-Revised (OCI-R) and Standardised Assessment of Personality – Abbreviated Scale (SAPAS) questionnaires were distributed to participants at baseline, but not at 18 months. OCI-R detects cases of Obsessive-Compulsive Disorder with an optimal cut-off point ≥ 21 for sensitivity and specificity of up to 0.74 and 0.75, respectively^24^. The SAPAS is validated in detecting cases of personality disorders in clinical populations with a sensitivity of 0.94 and specificity of 0.85 for a cut-off point of ≥ 3^25, 26^However, it is less accurate in the general population with a sensitivity of 0.69 and specificity of 0.53 at a cut-off point of ≥ 4^25, 26^.

### Demographic and clinical factors

At baseline, demographic data were collected, including age, gender, and biological sex. Ethnicity data were collected in UK census categories, and whether someone had an ethnicity making them vulnerable to hospitalisation from COVID was coded as ‘yes’ or ‘no’, based on the British Office for National Statistics reports of data accumulated from January 2020 to December 2021^16^. Those from South Asian, Black or ‘Other’ ethnic groups were deemed more vulnerable to COVID based on epidemiological data collected during the first wave of the pandemic^16^. Whether individuals had a medical condition that put them at a higher risk of hospitalisation from COVID was coded by a physician as ‘yes’ or ‘no’, based on the QCovid Risk Assessment model used by NHS England to classify these individuals’ self-reported health conditions^27^. At baseline, information about whether someone had a close family member hospitalised due to COVID was self-reported. Employment status was coded into categories: employed, self-employed, furloughed, unemployed, student and other. Data on household composition was obtained to indicate whether a participant lived alone or with others, and whether anyone in the house was perceived as being vulnerable to the effects of COVID-19. Finally, at the outset of the study, and at the 18-month timepoint, participants self-reported any previous hospitalisations due to COVID, previous COVID infection, and whether a participant had been vaccinated against COVID.

#### Data analysis

Statistical Package for the Social Sciences (SPSS) version 29.0.0.0 was employed to analyse study data, and GraphPad Prism 2.2.0 to create figures. Questionnaires required that each item was completed before the participants could move on to the next set of questions, ensuring that there was no within-instrument missing data.

Descriptive statistics were used to summarise the characteristics of the study participants. Data were assessed for normality by visually inspecting normal distribution curves for each variable and evaluating Kolmogorov-Smirnov and Shapiro-Wilk significance levels. The mean PHQ-15 score pre-pandemic was 7.97 (s.d. = 4.96, skew = 0.45, kurtosis = −0.42). The mean PHQ-15 score post-pandemic was 11.34 (s.d. = 5.72, skew = 0.18, kurtosis = −0.27). Differences between total PHQ- 15 scores and the differences between individual item scores were analysed using the Wilcoxon signed-rank test. As changes in PHQ-15 scores across all characteristic factors likewise did not follow normal distribution patterns, the Mann-Whitney U test was required for the statistical testing of these variables, except for age which was evaluated using the Kruskal-Wallis test. Spearman’s correlations were used to assess the correlation coefficient and significance level between PHQ-15 change and all psychopathology scales (baseline, 18 months and change in score). Multiple linear regression was utilised to explore factors associated with changes in the PHQ-15 score among study participants. Factors showing an association (p<0.2) in univariate analysis were retained in the multivariate model and entered using backwards step-wise selection^28^.

## Results

197 people provided complete survey data at 18 months follow-up. The average age of respondents was 40.9 (range 18 – 81). 159 (82.0%) of the sample were female, 32 (16.5%) male, and 3 (1.5%) of other gender. Regarding ethnicity, 141 (71.9%) reported being White British or Irish, 22 (11.2%) White Other, 12 (6.1%) South Asian, 2 (1.0%) Chinese, 8 (4.1%) Black, 4 (2.0%) mixed, 5 (2.6%) of another unspecified ethnicity and (2) 1.0% did not report their ethnicity. 29 (14.9%) reported being from an ethnic background that had a higher risk of becoming seriously ill with COVID-19.

At baseline, 89 (45.4%) of participants were employed or self-employed, 5 (2.6%) furloughed, 29 (14.8%) unemployed, 30 (15.3%) in education and 43 (21.9%) reported ‘other’. 15.3% reported living alone, while the rest reported living with a partner, family or friends.

By 18 months, 124 (62.9%) participants reported previously having had COVID, and 9 (4.6%) reported having previously been hospitalised with COVID. 48 (24.4%) participants reported they have a health condition which put them at higher risk of getting hospitalised with COVID (the most common were asthma, diabetes mellitus and rheumatoid arthritis) and 70 participants (35.5%) were living with or caring for someone vulnerable to getting seriously unwell with COVID^14^. 175 (88.8%) had received at least one COVID vaccine.

Cohort changes in mental health

Among this cohort of people recruited with severe COVID anxiety during the pandemic, there were significant improvements in all measures of mental health from baseline to the 18-month follow-up point. Whole cohort mean COVID anxiety reduced by 69.8%, with almost one in three no longer reporting any symptoms of COVID anxiety. Scores measuring generalised anxiety decreased by 31.7%, health anxiety by 23.9% and depressive symptoms by 24.5%. These changes are reported in greater detail in another analysis of this dataset^29^.

### Somatic symptoms before and after the pandemic

Somatic symptoms were commonly reported before the pandemic with 94.9% reporting at least one somatic complaint. The mean and median baseline PHQ-15 scores were 7.97(s.d. 4.96) and 8.00 respectively. The most common pre-pandemic symptoms were feeling tired or having low energy (67.5%), trouble sleeping (67.0%) and back pain (56.8%).

### Changes in somatic symptoms

The mean score reported by study participants was 7.97 (s.d. 4.96) before the pandemic and 11.34 (s.d. 5.72) after the pandemic. The increase in total PHQ-15 score was statistically significant (t = 9.043, p<0.001). Differences in the proportion of participants who had low, medium and high scores on the PHQ-15 are presented in Table 1.

**Table 1.** Changes in severity of somatic symptoms reported by 197 study participants.

| PHQ-15 Severity | Pre-pandemic (%) | Post-pandemic (%) | Percentage change (%) |
| --- | --- | --- | --- |
| Minimal (<5) | 27.9 | 12.7 | - 54.4 |
| Low (5-9) | 38.1 | 24.9 | - 34.7 |
| Medium (10-14) | 20.8 | 29.4 | + 41.4 |
| High (15-30) | 13.2 | 33.0 | + 150.0 |

To illustrate changes in scores of individual PHQ-15 items, mean individual item scores were calculated pre-pandemic and post-pandemic and plotted in Figure 1.

**Figure 1.**
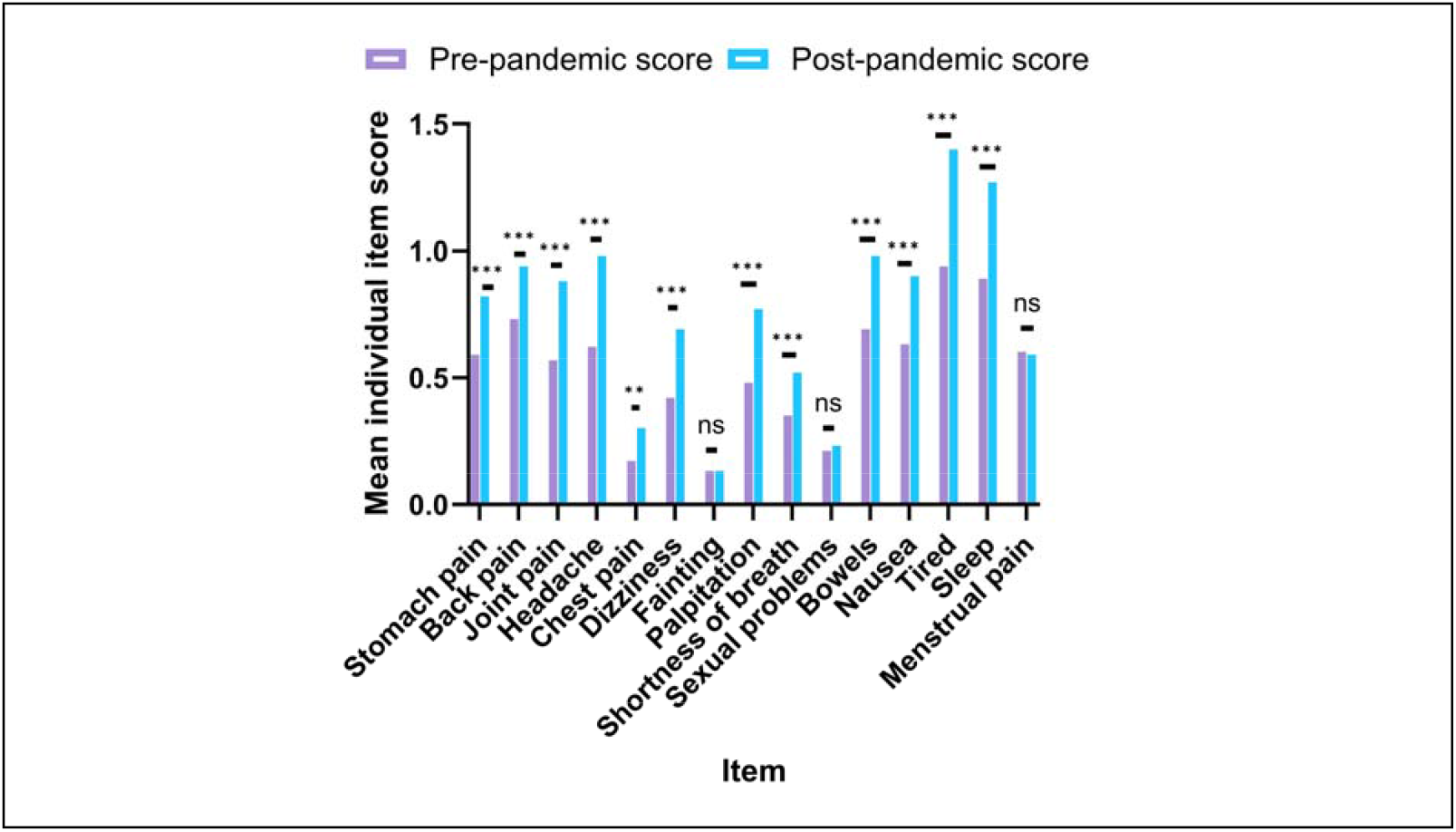
Histogram comparing mean individual PHQ-15 item scores pre-pandemic and post-pandemic. To explore changes in scores of individual PHQ-15 items, scores were compared before and after the pandemic using the Wilcoxon Signed-Rank Test. However, for most items the median score was zero or one. For illustrative purposes, mean scores were calculated and plotted in the histogram above. *p<0.05, **p<0.01, ***p<0.001, ns (not significant). Item titles are described in more detail in table 2. n=197

Change in score for each item, along with the percentage change and significance testing are presented in Table 2. The three items with the highest mean percentage change in score are: chest pain (+76.5%, p=0.002), dizziness (+64.3%, p<0.001) and palpitations (+60.4%, p<0.001). There were no apparent changes in fainting episodes, sexual problems or menstrual pain.

**Table 2.** The mean PHQ-15 individual item scores and the change in scores from pre-pandemic to post-pandemic. The Wilcoxon Signed-Rank Test was used to statistically assess changes in scores from pre-pandemic to post-pandemic. p-values are described in the fifth column and the percentage change is described as a percentage increase or decrease in mean from pre-pandemic to post-pandemic. All items had a statistically significant change in score except for fainting, pain or problems during sexual intercourse and menstrual cramps or other problems with period. n = 197.

| PHQ-15 Item | Mean pre-pandemic item score | Mean post-pandemic item score | Mean PHQ-15 item score change | Significance test (p=) | Standardised test statistic | Percentage change (%) |
| --- | --- | --- | --- | --- | --- | --- |
| Total score | 7.97 | 11.34 | 3.37 | <0.001 | 9.04 | 42.3 |
| Stomach pain | 0.59 | 0.82 | 0.23 | <0.001 | 4.59 | 39.0 |
| Back pain | 0.73 | 0.94 | 0.21 | <0.001 | 4.19 | 28.8 |
| Pain in arms, legs or joints | 0.57 | 0.88 | 0.31 | <0.001 | 5.59 | 54.4 |
| Headache | 0.62 | 0.98 | 0.36 | <0.001 | 6.03 | 58.1 |
| Chest pain | 0.17 | 0.30 | 0.13 | 0.002 | 3.17 | 76.5 |
| Dizziness | 0.42 | 0.69 | 0.27 | <0.001 | 5.42 | 64.3 |
| Fainting | 0.13 | 0.13 | 0.00 | 0.808 | 0.243 | 0.00 |
| Palpitation | 0.48 | 0.77 | 0.29 | <0.001 | 5.37 | 60.4 |
| Shortness of breath (SOB) | 0.35 | 0.52 | 0.17 | <0.001 | 3.60 | 48.6 |
| Pain or problems during sexual intercourse | 0.21 | 0.23 | 0.02 | 0.577 | 0.56 | 9.50 |
| Constipation, loose bowels or diarrhoea | 0.69 | 0.98 | 0.29 | <0.001 | 5.66 | 42.0 |
| Nausea, gas or indigestion | 0.63 | 0.90 | 0.27 | <0.001 | 5.21 | 42.9 |
| Feeling tired or having low energy | 0.94 | 1.40 | 0.46 | <0.001 | 6.59 | 48.9 |
| Trouble sleeping | 0.89 | 1.27 | 0.38 | <0.001 | 6.03 | 42.7 |
| Menstrual cramps or other problems with period | 0.60 | 0.59 | -0.01 | 0.851 | 0.19 | -1.70 |

Mann-Whitney U tests were used to determine whether changes in PHQ-15 scores were associated with demographic and clinical factors (see table 3). An ordinal categorical variable for age was created: 25 and below, 26-39, 40-59 and above 60. The Kruskal-Wallis test was used to establish if there was a significant difference between age categories.

**Table 3.**
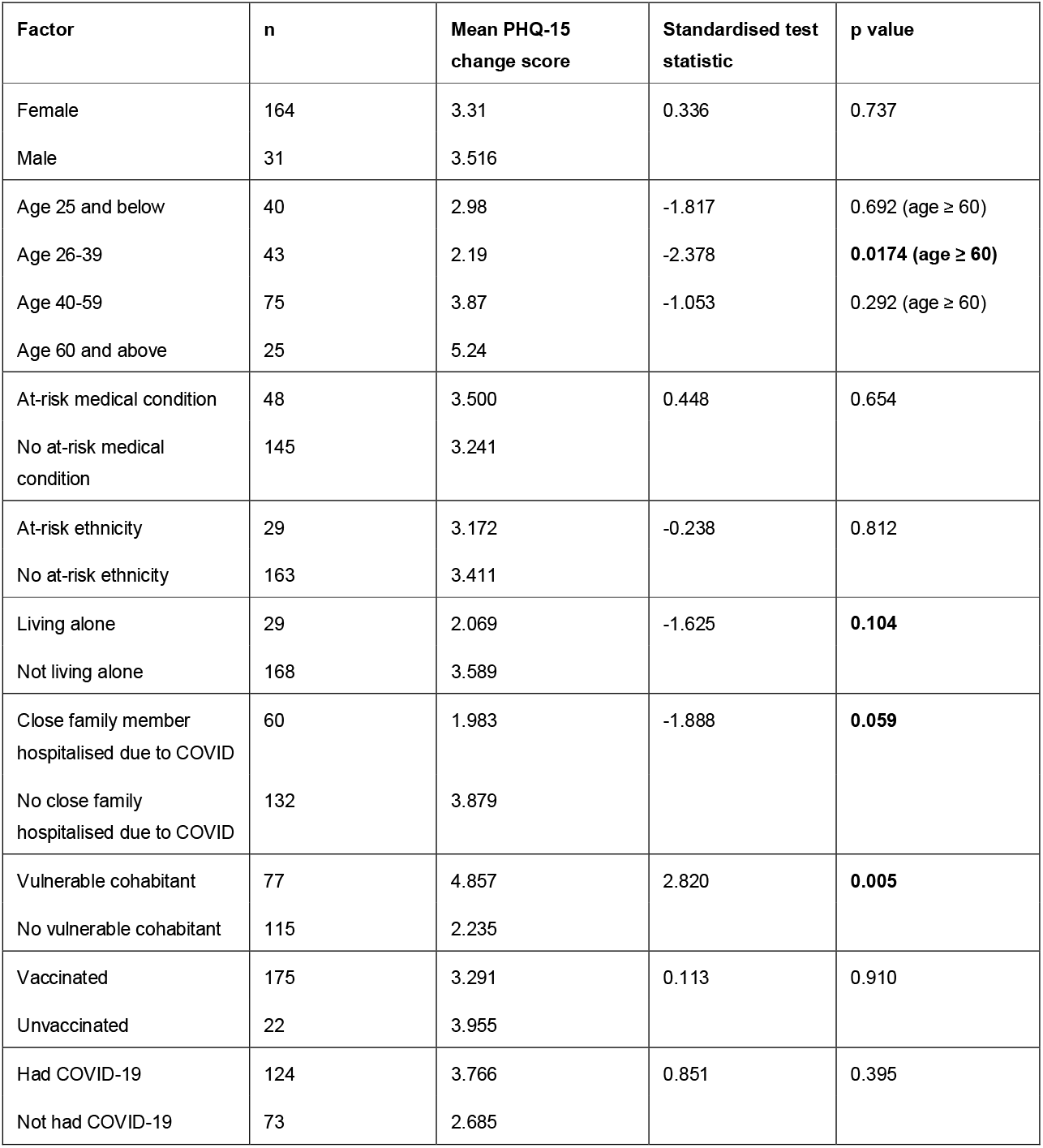
Describes a list of demographic and clinical factors and the mean and median change in PHQ-15 total score for each factor. The Mann-Whitney U Test was used to calculate the p-values for all factors except age, where p-values were calculated using the Kruskal-Wallis Test. The table shows p-values to indicate the significance of the change across each factor. The p-values highlighted in bold were < 0.2 and, therefore, these factors are included in the multivariate analysis. Total PHQ-15 scores for age group 26-39 were significantly different (p=0.0174) from those in age group 60 and above. Total PHQ-15 scores for those living with a vulnerable cohabitant were significantly different (p=0.005) than those living without.

People aged 60 and above had significantly greater changes in PHQ-15 scores compared to people aged 26-39. Those living with someone vulnerable to COVID also showed significantly greater changes in PHQ-15 scores compared to those who were not living with a vulnerable cohabitant.

### Correlations between psychopathology scales and change in somatic symptoms

Correlation coefficients were established for the psychopathology scales at baseline, 18 months and change in scale over the 18-month period, as shown in Table 4.

**Table 4.** Demonstrates correlation coefficients for PHQ-15 pre-pandemic score, post-pandemic score and change in score compared to psychopathology scale scores at baseline and 18 months. These were calculated using Spearman’s Rank Correlations with p-values indicated in the brackets on the right of the coefficient. The changes in CAS, GAD-7, sHAI-14, AUDIT-C and PHQ-9 all showed a significant correlation (p<0.05) with change in PHQ-15 score.

| Score | Pre-pandemic PHQ-15 Score | Post-pandemic PHQ-15 Score | Change in PHQ-15 Score Pre/post Pandemic |
| --- | --- | --- | --- |
| CAS baseline | 0.043 (0.550) | 0.114 (0.111) | 0.044(0.538) |
| Change in CAS (baseline -18 months) | 0.218 (0.002) | 0.377 (<0.001) | 0.226(0.001) |
| GAD-7 baseline | 0.013 (0.862) | 0.196 (0.006) | 0.171 (0.018) |
| Change in GAD-7 (baseline – 18 months) | 0.348(<0.001) | 0.532 (<0.001) | 0.316 (<0.001) |
| sHAI-14 baseline | 0.285 (<0.001) | 0.354 (<0.001) | 0.118 (0.105) |
| Change in sHAI-14 (baseline – 18 months) | 0.114 (0.117) | 0.318(<0.001) | 0.280 (<0.001) |
| AUDIT-C baseline | -0.040 (0.585) | -0.085 (0.247) | -0.056 (0.442) |
| Change in AUDIT-C (baseline – 18 months) | -0.064 (0.384) | -0.191 (0.009) | -0.157 (0.031) |
| PHQ-9 baseline | 0.141 (0.052) | 0.280 (<0.001) | 0.135 (0.063) |

Associations between mental health scale scores and changes in somatic symptoms are presented in Table 5.

**Table 5.** Multiple linear regression model showing coefficient β (beta), standard error, t score, significance (p-value) and confidence intervals. Significant values of p < 0.05 are highlighted in bold. Adjusted R^2^ = 0.137. Living with a vulnerable cohabitant, GAD-7 change were associated with increases in the PHQ-15 score (p=0.0273, p=0.0296 respectively). Change in AUDIT-C was associated with decreases in the PHQ-15 score (p=0.0452).

| | Coefficient $\beta$ | Coefficients standard error | t score | 95% Confidence intervals | Significance test ($p=$) |
| --- | --- | --- | --- | --- | --- |
| Constant | 4.00 | 1.41 | 2.82 | 1.21-6.80 | **0.0053** |
| Vulnerable cohabitant | 1.79 | 0.802 | 2.23 | 0.202-3.37 | **0.0273** |
| Close family member hospitalised | -1.44 | 0.759 | -1.90 | -2.94-0.0597 | 0.0598 |
| Living alone | -1.50 | 1.07 | -1.40 | -3.61-0.610 | 0.162 |
| Age | 0.0209 | 0.0245 | 0.851 | -0.0275-0.0692 | 0.396 |
| GAD-7 change | 0.189 | 0.0862 | 2.19 | 0.0189-0.359 | **0.0296** |
| sHAI-14 change | 0.0335 | 0.0617 | 0.543 | -0.0882-0.155 | 0.588 |
| PHQ-9 change | -0.0105 | 0.0741 | -0.141 | -0.157-0.136 | 0.888 |
| AUDIT-C change | -0.402 | 0.199 | -2.02 | -0.795-0.00861 | **0.0452** |
| CAS change | 0.0628 | 0.0964 | 0.651 | -0.128-0.0253 | 0.516 |

Multiple linear regression was utilised to explore factors associated with changes in PHQ-15 score before and after the pandemic. Two factors were found to be associated with an increase in somatic symptoms: greater increases in generalised anxiety and living with a person vulnerable to hospitalisation from COVID. Increased consumption of alcohol over time was found to be associated with decreases in somatic symptoms. The model explained 13.7% of variance in changes in the level of somatic symptoms.

## Discussion

This study describes changes in somatic symptoms reported among people with severe COVID anxiety before and after the coronavirus pandemic. During this period, the proportion of people reporting a high level of somatic symptoms more than doubled, from 13% to 33%, and the proportion of people experiencing no or minimal somatic symptoms more than halved. There was a clinically and statistically significant increase in somatic symptoms for almost all the individual items of the PHQ-15, except for ‘fainting’, ‘pain or problems during sexual intercourse’ and ‘menstrual cramps, or other problems with period’. The three symptoms with the highest percentage increase in score were chest pain, dizziness, and palpitations. This was observed despite major cohort reductions in other scores of poor mental health after the pandemic.

When adjusting for other demographic and clinical factors, those with severe COVID anxiety who lived with someone vulnerable to COVID-19, and who reported increasing levels of generalised anxiety during the pandemic reported greater increases in somatic symptoms. Increasing alcohol consumption was associated with a small decrease in reports of somatic symptoms (p=0.0452). Other factors like age, gender, having had COVID-19, vaccination status, or changes in other mental health scores like health anxiety or depressive symptoms did not predict change.

In a previous study of the general UK population, mean PHQ-15 score during the pandemic was 3.7, whereas, in the present study, the mean PHQ-15 score was 11.34 after the pandemic^10^. Comparatively the present study supports the previous findings of Shevlin et al., that after controlling for generalised anxiety disorder, moderate and higher levels of COVID anxiety are associated with greater rates of somatic symptoms^10^, and that this cohort with severe COVID anxiety have correspondingly higher somatic symptoms than the general population.

Among the general adult UK population gastrointestinal and fatigue symptoms^10^ were reportedly the most associated with increasing COVID anxiety. While these symptoms were reported in magnitude among our cohort of people with severe COVID anxiety too, we observed that the highest percentage increases were in symptoms that are well-documented to be associated with panic disorder (chest pain, dizziness, and palpitations)^30^. Whilst this study did not include a direct measure of panic disorder, it highlights that people with severe COVID anxiety may be more likely to experience symptoms similar to those of panic disorder^31^.

This may be bidirectional: research investigating Covid Stress Scale scores in anxiety and mood disorders demonstrated higher scores in those with panic disorder^32^. In addition to an increased number of google searches for panic-related symptoms^33^, there is previous evidence to suggest that during the pandemic across 11 countries, including the UK, there was an increase in these somatic panic disorder symptoms^34^. Predictors of developing a panic disorder during the pandemic were an increase in stress, pre-existing mental disorder, fear of infection, and the restrictiveness of the measures imposed by the government during the pandemic^34^. This cohort is more fearful of infection than the general population and, therefore, may be at a higher risk of developing a panic disorder during the pandemic, although further research is needed to establish this. It has been previously suggested that two of the symptoms in the CAS are the same as those experienced in panic disorder: feeling unable to move and dizziness^35^, which brings the question of whether using high CAS scores as inclusion criteria pre-selects individuals with acute anxiety and panic.

In contrast to our hypotheses, changes in CAS score were not associated with changes in somatic symptoms, but this may be attributed to the ‘ceiling effect’ of the CAS^36^. This effect occurs when questionnaire scores are initially high, limiting the potential explanatory power of further increases^36^.

Previous studies have suggested that people with higher generalised anxiety symptoms are more likely to experience somatic symptoms^10, 37, 38^, this present study adds the findings that increasing GAD-7 scores had a positive association with reported increases in somatic symptoms among people who had severe levels of COVID anxiety. There is a hypothesis therefore that alleviating anxiety may reduce levels of somatic symptoms according to a study assessing the impact of a behavioural medicine intervention in the US^39^ This could be particularly useful in the context of people who are highly anxious about COVID, as targeted interventions could be useful in decreasing both their anxiety levels and their somatic symptoms.

Living with someone vulnerable to COVID was also a significant positive predictor of PHQ-15 change in the multiple linear regression model. A recent study has highlighted an association between living with someone at high-risk of severe COVID and worsening scores in anxiety, depression, and the COVID-19 Stress Scale^40^. These individuals may have good reason for a heightened awareness of any signs of illness as potential symptoms of COVID, with corresponding increases in self-monitoring and bodily-vigilance due to heightened concern for the consequences of themselves and their co-habitant contracting COVID. Additionally, this subgroup of people living with someone vulnerable to COVID could represent a large proportion of carers. Some research into this specific group of people identified increased social isolation both during the pandemic, and universally in comparison to the general population^41^, social-isolation being a potentially confounding factor given that a recent study has shown that social isolation and fear of COVID are both directly related to somatic symptoms, and further act on somatic symptoms through a mediating factor of perceived anxiety^42^. ‘Living alone’, and other household arrangements reported by respondents did not show an association with change in the multiple linear regression model, but living alone does not necessarily indicate social isolation.

One unexpected finding of this study is that self-reported increases in alcohol use were associated with a decrease in somatic symptoms. This supports results from a study from 2015 in Norway, which found that people who abstained from alcohol showed higher levels of somatic symptoms than those who drank any amount of alcohol^43^. However, several studies have reported substance use and heavy drinking being associated with higher levels of somatic symptoms, so the association remains unclear^44^.

Finally, it is now well-established that COVID-19 can produce long-term somatic symptoms, and it is reported that as many as 1 in 8 COVID patients have persistent somatic symptoms^45^. Although PHQ-15 scores were overall higher in people who had previously had COVID, no significant difference was detected. This is in contrast with previous research, such as a study in South Korea, which found that people who had experienced COVID were more likely to have higher scores on the PHQ-15 than those who had not contracted COVID^46^. It is possible that this study was not sufficiently powered to detect a significant difference, as the symptoms of long-COVID are well documented in the literature^47, 48^.

### Strengths and Limitations

This study is the first exploration of how somatic symptoms evolved among people who experienced high levels of anxiety about COVID during the pandemic. It provides insight into the factors that may influence the change in somatic symptoms people may experience during a pandemic. The PHQ-15, CAS and psychopathology scales are all validated measures which can be useful in comparing results across studies and enhancing reproducibility. A further strength is that the study includes longitudinal data, which enabled the assessment of changes in measures over time, compared with existing cross-sectional reports^10^.

These data and analyses are limited in several ways, firstly, despite the option for participants to complete interviews by telephone this was rarely used, recruitment predominantly via social media can introduce a bias towards individuals who have some digital literacy and can exclude people who may not be as comfortable with or have access to technology. Ofcom’s digital exclusion review (2022) suggests that those with a condition limiting their ability to use technology, people with lower incomes, unemployed, older or living alone are more likely to be vulnerable to digital isolation^49^. Therefore, the sample may not be representative of the true population of people who are anxious about COVID.

Another limitation of the study was that the measure of somatic symptoms experienced by participants prior to the start of the pandemic was retrospectively reported. The PHQ-15 has been validated as a contemporaneous measure of somatic symptoms, but has not been used to capture somatic symptoms that people have experienced in the past^50^. Ideally, data from a cohort study that had assessed the prevalence of somatic symptoms before and after the start of the pandemic should have been used, but such a cohort does not exist. Asking people about symptoms before the pandemic may have introduced recall bias, in which people’s current health status affected their responses to questions about the presence or absence of somatic symptoms prior to the start of the pandemic. Finally, the CAP study was designed to assess changes in the level of COVID anxiety that people experienced during the pandemic, and it was not powered to assess predictors of changes in somatic symptoms. It is possible that the sample size available for this analysis was not large enough to identify factors that could have been associated with increases in somatic symptoms, such as whether people who had COVID had increases in somatic symptoms.

### Future perspectives

Overall, the results suggest that since the end of the pandemic reports of somatic symptoms have increased among individuals who had been severely anxious about COVID-19. People who have co-occurring higher levels of generalised anxiety or were living with someone vulnerable to COVID- 19 may be particularly vulnerable, here implicating a complex psycho-social process associating COVID anxiety and somatic symptoms. Further research is required to clarify the role of other complex social mediating factors, such as socialisation. Interestingly, research by another group during the COVID pandemic has shown via psychological network analysis and structural equation modelling that the cognitive-behavioural model (CBM) may also explain somatic symptoms^51^. State negative affectivity was found to predispose people to experiencing somatic symptoms during the pandemic^51^. Correspondingly some evidence links negative affectivity with COVID anxiety.

Together this raises the hypothesis that people with high trait neuroticism/negative affectivity may be predisposed to developing pandemic anxiety, and subsequently precipitate somatic symptoms^51^, by priming bodily hypervigilance, avoidance and safety-seeking behaviours, attentional biases and unhelpful changes in predictive coding.

The findings of this report also raise the possibility that targeting generalised anxiety and panic symptomatology or supporting those living with someone vulnerable could have an impact on reducing the substantial increase in somatic symptoms observed. Research has already shown that some interventions, particularly Cognitive Behavioural Therapy techniques may be helpful for improving anxious and depressive symptomatology among people with COVID anxiety, but the impact on somatic symptoms does not appear to have been studied^52-54^.

From a public health perspective, it may be helpful to assess whether increasing social support, particularly for carers, during the later stages of a pandemic can assist with alleviating somatic symptoms in highly anxious groups. As internet searches for panic-related symptoms increased near the beginning of the pandemic, it may be useful to better signpost people to resources and helplines through online search engines^33^.

## Conclusion

This is the first study to assess somatic symptom changes in people who were severely anxious about COVID-19 during the pandemic. In this cohort, somatic symptoms increased substantially overtime despite significant cohort reductions in other mental health symptoms after the pandemic ended. Particular attention should be paid to those living with someone vulnerable to COVID. Given that greater levels of somatic symptoms are associated with a poorer quality of life this finding is notable, and warrants further clinical attention since the end of the pandemic.

## Data Availability

The data used during the current study are available from the corresponding author on reasonable request.

## Acknowledgements

Authors have no acknowledgments to make.

## Notes

Funding statement: This research is supported by funding from the NIHR Imperial Biomedical Research Centre (WBPP_PA2902), a National Institute for Health Research Health Technology Assessment Programme (16/157/02), and an NIHR Senior Investigator Application held by Professor Crawford (NF-SI-0515–10006). The views expressed are those of the authors and not necessarily those of the NIHR or the Department of Health and Social Care. The sponsor of the study is Imperial College London.

### Competing Interest Statement

The authors have declared no competing interest.

### Clinical Protocols

https://bmjopen.bmj.com/content/12/9/e059321.long

### Funding Statement

This research is supported by funding from the NIHR Imperial Biomedical Research Centre (WBPP_PA2902), a National Institute for Health Research Health Technology Assessment Programme (16/157/02), and an NIHR Senior Investigator Application held by Professor Crawford (NF-SI-0515-10006). The views expressed are those of the authors and not necessarily those of the NIHR or the Department of Health and Social Care. The sponsor of the study is Imperial College London.

### Author Declarations

Leicester Central Research Ethics Committee and Health Regulation Authority approved the COVID Anxiety Project, reference number 20/EM/023. The study adhered to the guidelines established by the Declaration of Helsinki 1964. Data were handled according to the General Data Protection regulations, and Qualtrics is certified under ISO 27001, ensuring it adheres to international standards for handling sensitive and confidential information. All participants signed a consent form before taking part in the study. Once completed, the consent forms were printed and physically filed securely. Participants could withdraw from the study at any point with no explanation required.

